# Factors influencing N95 respirator use among healthcare workers in a TB specialized hospital, Bangladesh

**DOI:** 10.1101/2025.09.01.25334196

**Authors:** Sayeeda Tarannum, Abu-Hena Mostofa Kamal, Mohammad Tauhidul Islam, Md. Ariful Islam, Kamal Ibne Amin Chowdhury, Shirina Akhter, Afsana Sharmin, Emily S Gurley, Sayera Banu, Md. Saiful Islam

## Abstract

**Background:** Tuberculosis (TB) infection prevention and control (IPC) guidelines recommend that healthcare workers (HCWs) wear N95 respirators in pulmonary TB patient areas. To ensure appropriate use of respirators by HCWs, an understanding of the barriers and facilitators associated with respiratory protection is needed. This study explored factors influencing N95 respirator use among HCWs in a TB-specialized, tertiary-care hospital in Bangladesh.

**Methods:** Between February 1, 2014 and March 14, 2014, a field team consisting of five social scientists and three epidemiologists conducted this qualitative study in a tertiary-care, specialized TB hospital in Dhaka, Bangladesh. Initially, we held a day long training workshop on respirator use, re-use, and storage, and conducted N95 respirator fit tests. After the workshop, we provided a two-month supply of N95 respirators for HCWs assigned to two multi-drug resistant (MDR) TB inpatient wards. Finally, we conducted in-depth interviews with the HCWs to explore their risk perceptions regarding TB and airborne infections, and reported advantages and disadvantages of N95 respirator uses. We used the HBM health belief model to understand the factors that influence respirator use.

**Results:** Doctors and ancillary workers reported that discomfort, demotivation from the senior colleagues, and cumbersome storage of respirators for re-use were the primary barriers to respirator use. In addition, communication difficulties, fogging up corrective lenses, eight hours of respirator use are considered factors that also impacted respirator use. However, compared to doctors and ancillary workers, the nurses are more likely to use N95 respirators.

**Conclusion:** Workshops on TB transmission and prevention strategies, and easy access to those strategies, might improve the use among the nurses. Interventions that target educating HCWs on TB transmission and the role of personal protective equipment in TB prevention, as well as hanging posters on the wards as reminders, supplying and ensuring easy access to N95 respirators inside the ward, and developing communications targeted at behavioral change, including advocacy and counseling, can also be implemented.

## Introduction

In low- and middle-income countries (LMICs) with high tuberculosis (TB) burdens, healthcare workers (HCWs) are at risk of acquiring TB infection due to repeated and sustained exposures to TB patients in healthcare settings without proper implementation of TB infection prevention and control (IPC) measures(1). To protect HCWs from occupational TB infection, the World Health Organization (WHO), US Centers for Disease Control and Prevention (CDC), and Bangladesh National TB Control Program (NTP) recommend the use of N95 respirators among HCWs as part of a multi-component strategy to transmission of *M. tuberculosis* in health care settings (2–4). The CDC and WHO also provide guidance on the re-use or extended use of respirators by HCWs in instances where respirators are in limited supply(5).

The Bangladesh NTP IPC guidelines have recommended the use of particulate (N95) respirators for HCWs when they care for patients with pulmonary TB (6). To ensure proper respirator use among HCWs in Bangladesh, an understanding of individual, administrative, and cultural issues associated with respiratory protection is needed. In this study, we provided training and N95 respirators to HCWs in a TB specialty, tertiary-care hospital in Bangladesh and subsequently assessed reported barriers and facilitators to HCWs’ respirator use.

## Materials and Methods

### Study site and participants

We conducted this study at a 685-bed, tertiary-care, TB specialty hospital in Dhaka, Bangladesh. The hospital provided services to approximately 100,000 TB and respiratory patients per year, referred from hospitals and specialized doctors throughout the country (7). All doctors, nurses, and ancillary workers (sweepers, ward servants, *ayas*) who worked on the hospital’s two multi-drug-resistant (MDR) TB wards were invited to participate.

### Study design and data collection

We implemented a TB IPC intervention consisting of providing training and N95 respirators to HCWs, followed by a qualitative study on barriers and facilitators to respirator use among the participating HCWs. A 7-member, field team consisting of social scientists and epidemiologists trained in qualitative research conducted the study. The team talked to the hospital director and the heads of the study wards to describe the study objectives, and obtain permission to conduct the study. After receiving permission, we conducted a training workshop on respirator use and fit testing. The biosafety and biosecurity team from the International Centre for Diarrhoeal Disease Research, Bangladesh (icddr,b) led the workshop and provided the training. The team described TB transmission pathways, the role of N95 respirators in preventing TB and other respiratory diseases. The experts also facilitated a hands-on training on the proper use, re-use, and storage of respirators. Participating HCWs also received respirator fit testing and were supplied with appropriately sized respirators. Zip-lock bags (we made two small holes in the zip-lock bag for ventilation using a punch machine) were supplied with the respirators to store respirators between uses. We provided a total of 171 National Institute for Occupational Safety and Health (NIOSH)-certified, N95 respirators (3M Corporate Headquarters 3M Center St. Paul, MN 55144-1000, USA) to 24 HCWs (mean 6.6 respirators/HCWs) for 62 days. We maintained a register to document the date and time that respirators were provided. We tracked use time for respirators and instructed HCWs to use a new respirator after eight hours of total use during the care of TB patients. If HCWs reported feeling uncomfortable reusing a respirator, they were allowed to take a new one. We also instructed them to discard used respirators in the designated waste receptacles located outside the wards.

We selected all HCWs purposively, who worked in the MDR-TB wards, having TB-related experiences and their roles in TB prevention. We conducted 24 semi-structured, in-depth interviews (IDIs) with the HCWs to understand the barriers and motivators for N95 respirator uses. The respondents had a variety of educational backgrounds. All the participants were native Bengali speakers, so the interviews were conducted in Bangla. We asked the respondents about the motivating factors, such as safety concerns and professional obligations, and demotivating factors, including technical issues, physiological barriers, risks of contamination, and socio-cultural barriers for using N95 respirators. We recorded the interviews using a digital audio recorder, and the average duration of the interviews was 42 minutes. The interviews were conducted at the workplace of the respondents suggested by the respondents. We also took detailed notes during interviews.

### Data analysis

A descriptive analysis was performed on the data obtained through the interviews. The research team transcribed the audio recordings verbatim in Bengali and then translated the content into English using Microsoft Word. The recordings were reviewed multiple times to ensure the accuracy of the transcriptions. Initially, two researchers examined the field notes to develop a preliminary codebook. The coding framework was informed by the study’s objectives, relevant literature, and both predetermined and emerging themes related to the motivations and challenges of N95 respirator use among HCWs. The transcribed content and field notes were imported into ATLAS.ti version 6.0 to facilitate coding and analysis. The lead researcher reviewed and coded relevant excerpts based on the established codebook. When new data did not fit the existing codes, new codes were developed and incorporated, and this iterative process continued throughout the analysis. Coders held regular discussions to refine and ensure consistency in code definitions and categorization of emerging themes. The coded data were organized under broader themes, and thematic summaries were developed in English. All members of the research team collaboratively reviewed the themes and sub-themes to reach consensus and ensure consistency. The analysis was guided by the Health Belief Model (HBM), which includes five key dimensions: (a) perceived susceptibility; (b) perceived severity; (c) perceived benefits; (d) perceived barriers; and (e) cues to action.

### Ethical consideration

We obtained informed, written consent from the hospital authorities for conducting the research activities. We obtained informed written consent from all IDI participants. The study protocol PR#12067 was reviewed and approved by icddr, b’s Institutional Review Board (IRB) consisting of a Research Review Committee (RRC) and an Ethical Review Committee (ERC).

## Results

A total of 28 HCWs attended the workshop: 12 doctors, nine nurses, and seven ancillary workers. Of the 28 healthcare staff working on the MDR-TB wards, all were invited to participate; however, we excluded one doctor and one nurse from fit-testing as both had beards, and a bearded person cannot achieve a full fit test (8). Another doctor and one ancillary worker were transferred to another department of the hospital and, therefore, excluded from the fit testing and interviews.

The mean age of the study participants was 40 years; were 13 (54 %) were female, nine (38 %) were nurses, 10 (41%) were doctors, and five (21 %) were ancillary staff. The average time working in a TB hospital ranged from 4.5 years for doctors to 16.5 years for ancillary workers. As a note, based on the fit testing, icddr,b’s expert found that 11 HCWs required regular-size size and the rest of the HCWs required small-size N95 respirators.

### Perceived need for N95 respirators among healthcare workers

Of the 10 participating doctors, three reported that they should use a respirator in the MDR-TB ward, the TB ward, and while examining suspected TB patients. Additionally, four of the doctors stated that the use of the respirator was needed if someone remains in close contact with any TB patient, especially in MDR-TB ward (Table 1).

**Table 1:**
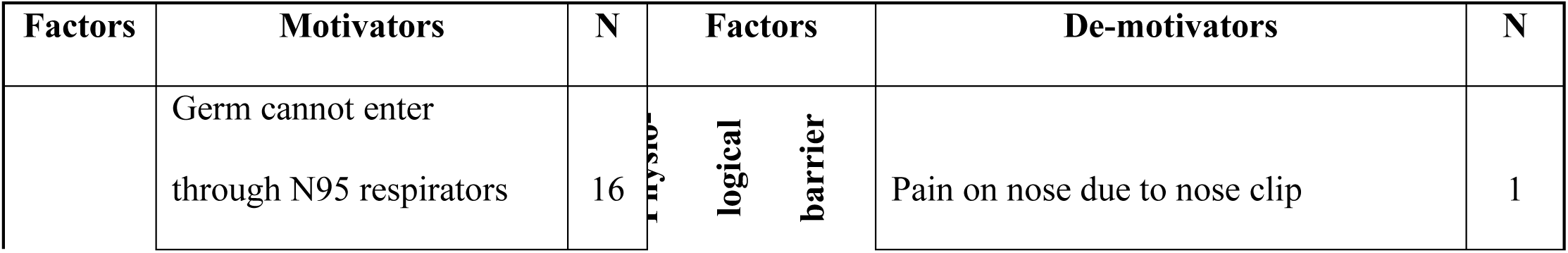
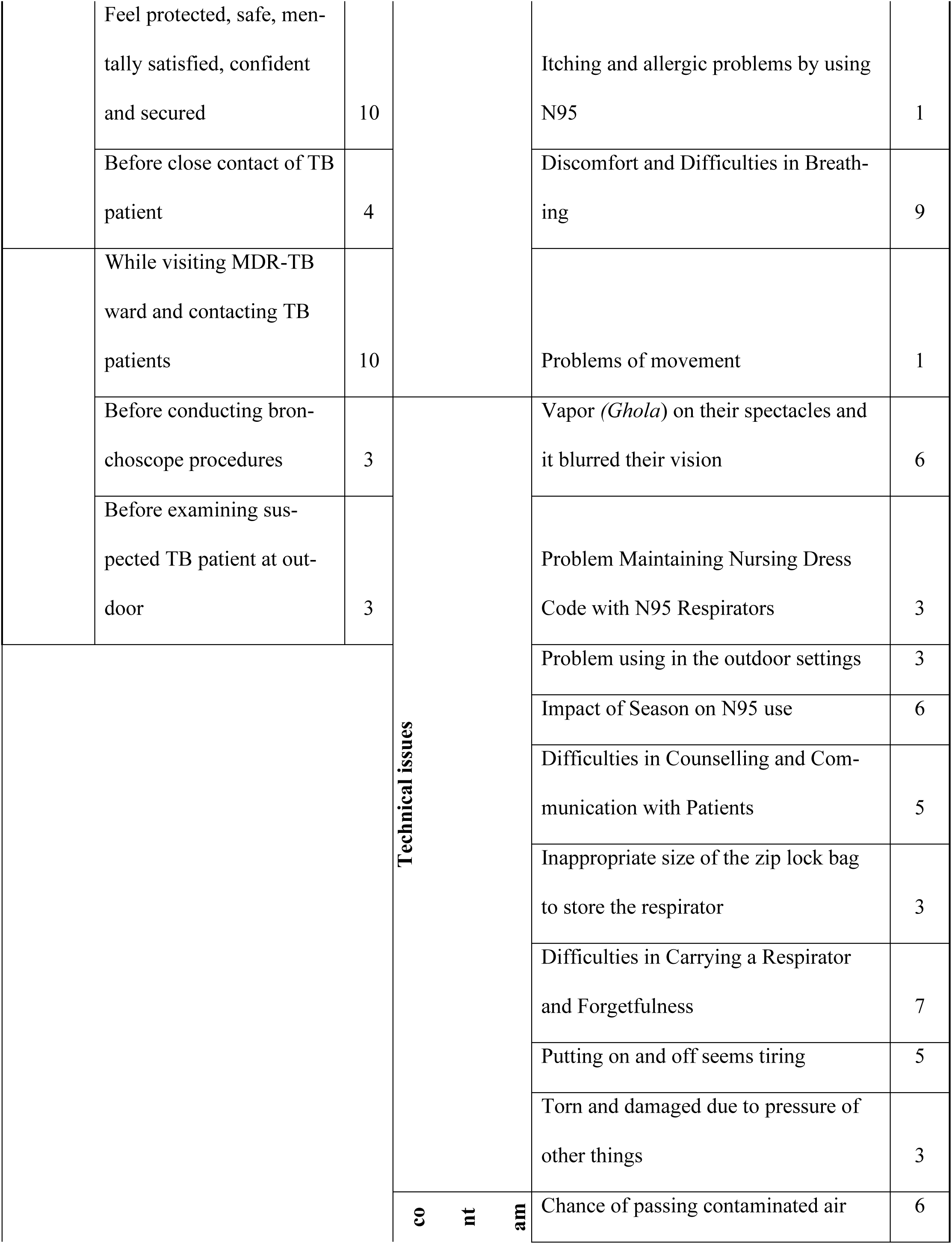
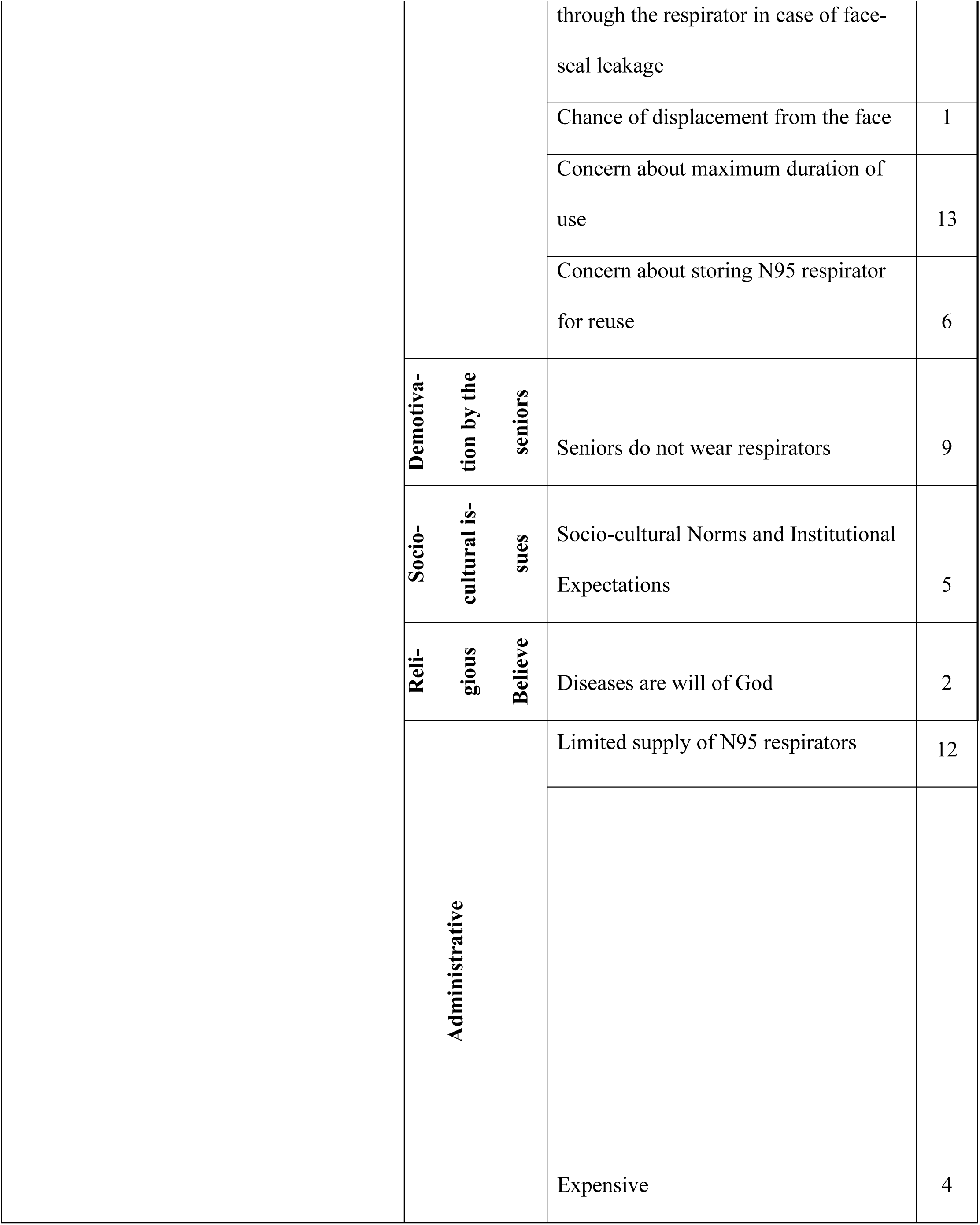
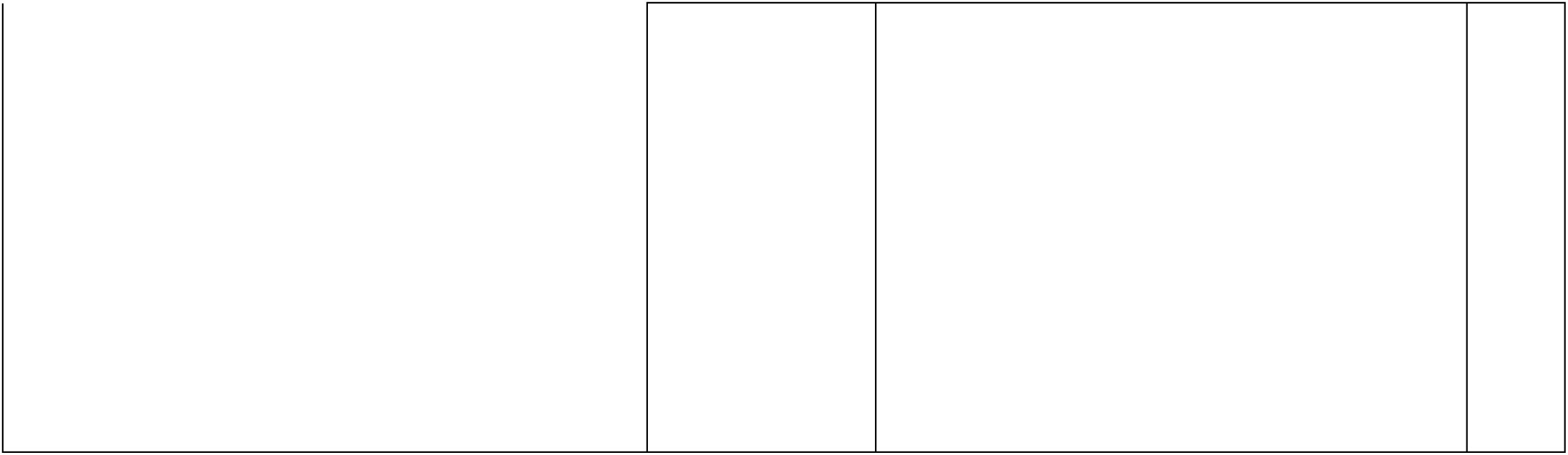
Factors influencing N95 respirator use among HCWs Working on MDR-TB Wards in a public, tertiary-care hospital, Bangladesh,2014.

### Healthcare workers’ perceived risk of TB infection

Of 24 participants, 18 (75%) perceived that they were at risk of TB infection due to close contact and everyday exposure to TB patients in the wards. One nurse stated,

> *I can be infected by the TB patients; I might get the infection from the patient’s coughing and sneezing. For this reason, I must always keep a distance from them. But at the same time, I must do my activities here when needed*.

Moreover, 7 (29%) participants mentioned that they were afraid of being infected with MDR-TB while providing care to patients. One nurse stated,

> *I will get TB if I talk to them. I will inhale their breath, they will inhale mine; All of us working in this hospital are at risk* [of TB infection]*. Staff, patients, and visitors are all at risk* – A male nurse.

One ancillary worker added that she was aware of the risk of being infected with TB, but she was not afraid, because she believed that “one gets sick from God’s will”. According to one of the nurses, “protection against TB was entirely up to God”.

On the other hand, one ancillary worker mentioned that he was not at risk of TB because he maintained cleanliness, washed his clothes regularly, and took a bath after returning home every day. A quarter of participants (6/24) informed us that they were concerned about getting TB infection during the initial days of beginning work at the TB hospital. Over time, they became accustomed to working with TB patients, and their concern about TB infection diminished.

In addition, one nurse and one doctor said that they heard from a senior colleague that all HCWs aged 40 and older are likely to have latent TB infection. Another doctor quoted from one of his senior colleagues that none of the doctors who worked at NIDCH were infected with TB, so he should not be worried about acquiring TB infection. Three participants provided an example that they observed very few HCWs working in the hospital develop active TB, so they felt confident in the low probability of being infected with TB. One doctor stated,

> *Our senior colleagues encourage us by saying they have been serving at this hospital for many years, and they have not been infected. So, we will not be affected. We have been working here with this belief* [we would not be infected].

### Factors influencing the use of N95 respirators

#### Perceived benefit of using N95 respirators

Sixty percent (16/24) of our study participants asserted that germs cannot enter through an N95 respirator; therefore, it can protect them from TB infection. Ten (42%) reported that they felt protected, safe, confident, and secure when they used an N95 respirator (Table 1). One doctor mentioned that an N95 respirator could protect against the TB germ if it is used appropriately. He said,

> *…the highest advantage is feeling safe. N95 respirator provides maximum protection against TB infection. I am feeling safe because I am wearing a respirator that will protect me from TB infection. Moreover, it gives me mental satisfaction and confidence since I am using respirators, I will not be infected*.

#### Cues to action

Nearly half of the participants (10/24) mentioned that they were concerned about MDR-TB, which worked as a cue to remind them to wear respirators before entering the ward. One of the doctors shared that TB patients’ exhale a large number of bacteria during bronchoscopy, as the procedure induces significant coughing. According to him, the bronchoscopy procedure reminds him to wear an N95 respirator. One nurse added,

> *All hospital staff who deal with TB patients emphasized wearing respirators. Everyone keeps in mind that they need to wear a mask before entering inpatient wards. Whenever we go to the inpatient ward or decide to go, we wear the respirator*.

#### Factors impeding the use of N95 respirator

Seventy percent of the doctors (7/10) said that they preferred inhaling fresh air to wearing a respirator. Furthermore, most of the doctors (6/10) perceived that the N95 respirator could ensure 95% protection against TB germs which denotes there always remain 5% chance of being infected despite wearing the respirator (Table 1). They also perceived that contaminated air could easily pass through the respirator if there is any face-seal leakage. One of the doctors mentioned,

> *It (respirator) does not remain fixed at the same position as we fit it (on face) at the beginning of wearing; it becomes loose and displaced. I feel uncomfortable when it becomes displaced. The germs may enter my respiratory tract due to loose fitting and displacement*.

#### Concern about N95 respirator storage and re-use

Few participants (3/24) believed that a used N95 respirator may be the source of TB infection if it is being reused and stored in the same zip-lock bag several times. One third of the doctors (3/10) added that carrying respirators either in a bag or inside their pocket was not safe or feasible. They added that respirators might get contaminated with other microorganisms in the zip-lock bag (Table 1). They also reported that they did not have any idea about how long a respirator could be stored in the bag and for how long it would remain safe for re-use. One doctor stated,

> *I do not want to use a respirator that I will have to carry in hand or in my pocket. It would have been better if we could store it in a backpack. It may increase the chance of infection if I keep it in my pocket and re-use it. I do not like keeping a used respirator in my pocket. If I do not properly store a respirator, I will not re-use it*.

#### Discomfort and difficulties in breathing

Nine (37.5%) study participants mentioned that a N95 respirator could not be used for greater than an hour constantly as it created difficulty breathing. One doctor said that an N95 respirator could be worn for 20-30 minutes at a time. The doctors reported feeling suffocated and difficulty breathing when using a respirator for an extended period of time. One ancillary worker stated that she wore respirators while transporting patients but could not continue after transporting the patient in the ward due to feelings of suffocation. All of the nurses (8/8) reported that it was not possible to wear a respirator throughout their entire duty hours; they only used the respirator while providing patient care on the ward. One doctor said she felt itchy when she used the respirator. One nurse reported that she had a feeling of pain on her nose when she used the respirators.

#### Impact of Climatic Conditions

A quarter of participants (6/24) mentioned that during the summer, facial sweat and retained exhaled breath generate additional heat on their face when they wear a respirator (Table 1). All participants stated that they must spend a long time with the patients; the average time worked by doctors and nurses was 6.5 hours and 8 hours per day (six days per week), respectively.

#### Difficulties in carrying a respirator and forgetfulness

Most of the study doctors (7 out of 10) reported that carrying a respirator from one ward to another was a barrier (Table 1). Since respirators were not available in the wards and doctors’ offices were located more than 100 meters away, they had to carry their respirators during ward rounds or while visiting patients. One doctor noted that the respirator felt like an additional burden during rounds, as she also had to carry student books and review patients’ investigation files, X-rays, and pathology reports.

Additionally, doctors frequently reported forgetting to bring their respirators during ward rounds. A few participants (3 out of 24) stated that their respirators became torn or damaged while being carried in a pocket or bag (Table 1). One ancillary worker mentioned that she did not use an N95 respirator while serving food to patients, as she needed to move quickly to distribute meals.

One nurse reported that sometimes in an emergency, for example if the patient’s condition became critical or if the visiting doctor on the ward called for an emergency, then the nurses had to run to the ward and sometimes forgot to wear a respirator.

#### Socio-cultural norms and institutional factors

Half of the study doctors (5/10) informed us that HCWs’ use of a respirator was not common in Bangladesh (Table 1). A doctor explained that wearing a gown is a standard precaution for doctors, however, very few HCWs comply with that standard, and similarly non-adherence to respirator use became a culture. He said,

> *The culture of self-protection has not been developed yet. For example -doctors should wear an apron, but we do not use it. Non-adherence has become a tradition or culture. This problem is due to a lack of habit.…many of us make fun of someone when they put on a mask*.

On the other hand, 3/8 nurses reported that there was a limited supply of respirators. They were instructed to use a respirator for seven days, but sometimes they had to use a respirator for two weeks owing to the limited supply. In addition, respirators were too expensive for HCWs to buy on their own. Each day the nurses had eight hours on duty and they stayed in close contact with patients on an average of three of those hours. Therefore, they reused the respirators for 3 hours a day for seven days a week or sometimes more than that. One nurse stated,

> *We almost stay the whole time with patients in the wards for taking care of the patients during ward rounds, providing injections to the patients. However, during the evening or night shift we spent less time but in morning we must stay longer hours with the patients*.

#### Nursing uniforms

Some nurses (3 out of 8) reported that their uniforms—particularly the nursing hat—posed a barrier to respirator use, as it prevented them from properly wearing the N95 respirator. One nurse in charge mentioned that nurses faced difficulties both putting on and removing respirators due to the hats. To address this, they removed their hats and instead wore cloth caps while working in the MDR-TB ward, where they needed to visit patients frequently.

One nurse explained:

> *“We are used to wearing a nursing hat as part of our uniform, and it makes us look beautiful. We had difficulties wearing an N95 respirator because of this large and specially designed hat. Now, we wear a small cloth cap so we can wear a respirator without any issues. See how much we are sacrificing to wear an N95 respirator.”*

#### Limited motivation from senior colleagues

Over half of the study doctors (6/10) mentioned that they followed the example of their senior colleagues regarding respirator use (Table 1). They hesitated, felt awkward, and remained confused about the necessity of N95 respirators because their senior colleagues did not wear them. One doctor said,

> *The main problem is that when you observe that your senior colleague is not using the respirators, you cannot use it. Suppose your professor is not using a respirators when he is visiting patients in the ward. In that case, how can I wear a respirator as a junior doctor?*

#### Difficulties in counselling and communicating with patients

Five of the study participants (5/24) reported that the use of a respirator was a barrier when communicating with patients. One doctor stated that she had to speak loudly while wearing a respirator. It also hampered their expressions and natural flow of speaking, she added. One nurse reported that the facility was an academic institute, and they had to communicate frequently with professors regarding patients’ conditions; the use of respirators became a barrier as it hindered the natural flow of conversation; therefore, they could not continue using a respirator (Table1). Six HCWs mentioned that their spectacles fogged up and blurred their vision when they used respirators.

## Discussion

This study found that the use of N95 respirators varied among healthcare workers (HCWs) and was challenging due to administrative issues, physiological discomfort, perceptions of contamination risk, supply constraints, lack of role models among senior staff, and sociocultural and technical barriers.

The supply of N95 respirators at the study hospital was limited, and HCWs expressed inconsistent beliefs about the effectiveness of respirators. Discomfort and breathing difficulties were commonly reported as barriers to use. In addition, concerns related to the storage and re-use of respirators, as well as a lack of motivation from senior colleagues, further discouraged use. Other reported challenges included the need to carry respirators between wards, forgetting to bring them, and difficulties in communicating with patients while wearing them.

Previous studies have similarly found that factors such as individual decision-making, motivation, risk perception, perceived disease severity, equipment availability and accessibility, monitoring and supervision, and cultural influences—including values, behavior, encouragement, and peer feedback—affect N95 respirator use (9–16).

In our study, nurses were found to be more consistent and frequent users of N95 respirators compared with doctors and ancillary staff. This may be attributed to their longer and more frequent exposure to TB patients, greater perceived risk, and easier access to respirators, which were stored near their duty stations adjacent to the study wards.

Overall, our findings suggest that simply providing N95 respirators may not be sufficient to ensure their consistent use. A comprehensive approach—addressing logistical, behavioral, and cultural factors—is essential to improving adherence to respiratory protection practices among HCWs.

Discomfort associated with respirator use was an issue impacting the acceptability. Our study also identified that a few of the HCWs were not adherent to respirators use due to breathing difficulties which was consistent with prior studies that reported discomfort associated with respirator use affected the adherence among HCWs in the in-patient wards in tropical countries (13, 15, 17, 18). In hot humid temperature, use of N95 respirators may increase concentration of CO2 and reduced concentration of O2 for inhalation and cause dizziness, nausea and headache (19, 20). HCWs felt comfortable wearing N95 respirator in summer with a temperature between 20 and 24 degrees Celsius and a relative humidity between 76%-96% (19, 21). It might be difficult to use respirators, particularly in weather like Bangladesh where the average annual temperature in summer time is found to be 26.9 to 31.1 degree Celsius (22)

During emergency, the use of a respirator among HCWs was low due to HCWs heavy involvement in setting critical patient and forgetfulness to take a respirator. The N95 respirators was stored at the doctors’ chambers or lounge which were a bit away from the study ward (23), which was found a significant barrier to comply with N95 respirator use (24). A study revealed that adherence is enhanced when Personal Protective Equipment are readily available (25).

On the other hand, the doctors were involved in providing care to multiple in-patients wards and the distance to doctors work station to collect a respirator was a barrier to respirator use (18). Thus one study revealed that the respirator use among the doctors could be increased by having an arrangement of storing respirators in a place nearby the patients’ ward (26). Therefore, an arrangement can be made by the hospital authority for storing the respirators for doctors in front of the patients’ ward.

Training on personal protective equipment among HCWs improves risk perception and possibly adherence (27). In our study, although we did not find HCWs perceived need of N95 respirators use, particularly among doctors and ancillary workers, training on TB transmission and respirators use had a positive impact in improving HCWs knowledge about TB risk perception as the HCWs mentioned in the interviews and how to re/use and store a respirator and the role of a respirator in TB prevention which are likely to improve the adherence if the other barriers are addressed (1, 10, 16, 26, 28, 29). The use of N95 respirators is key component of airborne infection prevention control and should be used in combination with administrative controls (e.g.,) and environmental controls (e.g., ventilation). A study recommended some alternatives, such as using an intubation box while doing the induction procedure, installing negative pressure machines in the rooms, or only allowing the essential healthcare workers in the room(33). However, a study conducted in the United Kingdom invented a technique named *singh thatta* technique, it is a under mask plastic made bearded cover for those who have beard. This technique was proven successful in achieving fit testing of the respirators(34).

Our study has limitations. First, this study in only one specialized hospital in Bangladesh and the findings may not be generalizable to other facilities participant selection. Second, this qualitative study relied on self-reported information and thus is subject to reporting bias. Our study is context-specific as we conducted this study in only one specialized hospital in Bangladesh and may not be generalizable to other facilities. Thus, the compliance rate might vary during summer due to sweating inside the respirator (35). However, the barriers and facilitators identified in this study are aligned with other recent studies.

These findings suggest that many of the barriers to HCWs respirator use could be addressed by maintaining adequate respirator supply, conducting routine respirator fit-testing, and regular training on N95 respirator use (36). Provision of respirators with an exhalation valve may help to address discomfort due to difficulty breathing and warm temperatures (37). Posters can be hung on doctors’ room or respirators can be made available near the wards entrance to improve adherence.

Additionally, having senior doctors IPC champions and role models may also increase adherence among junior doctors and other HCWs. Furthermore, placement of standardized signage outside the TB ward might be a useful reminder for the HCWs to wear N95 respirators (38, 39).

## Conclusion

This study found that N95 respirator use among HCWs was feasible, but the use varied by HCW groups, with The highest reported adherence being among nurses. Due to the limited supply of respirators in hospitals, discomfort, communication barriers, and lack of knowledge about reuse and storage, implementing and maintaining the recommended respiratory protection programs is challenging.

To improve adherence among all HCWs, interventions that target training HCWs, particularly doctors, and ancillary workers about TB risk and the role of a respirator in TB and other respiratory disease prevention should be implemented. Besides, TB IPC committee can be formed and the senior HCWs can be involved in TB IPC committees as IPC champions and opinion leaders and encouraged them to motivate juniors wearing N95 respirators. In addition, respirators should be made available inside the study wards and easily accessible to HCWs through ensuring regular supply.

## Data Availability

Data are available from the data repository committee at icddr,b. A copy of the complete dataset (anonymized and decoded) of this study will remain at the data repository. Interested researchers may contact Head of research administration, for approval and data access.

## Acknowledgement

We express our sincere gratitude to all the study participants and thank icddr,b’s core donors for unrestricted support. We are grateful to Dr. Michele L. Pearson, US Centers for Disease Control and Prevention, for his valuable input during manuscript writing. We are also thankful to Mr. Adib Shamsuddin, MD, BASc, Department of Family Medicine, University of Western Ontario, Chatham, Canada, for reviewing the manuscript and giving valuable suggestions.

